# Methylphenidate use and restorative treatment in 13,000 young adults with ADHD

**DOI:** 10.1101/2020.04.21.20074567

**Authors:** Haggai Schermann, Nathan Schiffmann, Ran Ankory, Amir Shlaifer, Nirit Yavnai, Victoria Yoffe, Lena Natapov

## Abstract

**Objective:** To assess a dose-response effect of methylphenidate use on restorative treatment needs, which served as an indicator of caries.

**Subjects and Methods:** This study is a retrospective cohort of military recruits aged 18-25 who served for 12 to 48 months between 2005 and 2017. The cohort included 6,875 subjects with ADHD who received treatment with methylphenidate, 6,729 subjects with ADHD who had no prescriptions for methylphenidate, and 200,000 healthy controls. The outcome was normative treatment needs: having at least one recommendation for restorative treatment during the study period.

**Results:** Frequency of recommendation for restorative treatment among the treated, the untreated and the control groups was 24%, 22% and 17%, respectively (p<0.0001). On multivariate analysis, the dose-response association between methylphenidate use and the odds of having at least one restorative treatment was confirmed (OR=1.006 for each additional 1 gr of methylphenidate; 95% CI [1.004:1.009]).

**Conclusions:** Subjects with ADHD who receive chronic treatment with methylphenidate have higher restorative treatment needs than subjects with untreated ADHD and healthy controls. Our results show that chronic methylphenidate medication among young adults leads to an elevated need for restorative treatment and implies a significant impact on oral health.

## Introduction

Methylphenidate (MP) is an effective stimulant medication for treatment of Attention Deficit and Hyperactivity Disorder (ADHD). Its use is prevalent in up to 7.5% of children of ages 6 to 18 and in up to 35% of college students (Ben-Ami et al., 2018). The research of the adverse effects of MP on patient health reported osteopenia (Feuer, Thai, Demmer, & Vogiatzi, 2016) and slower growth in children (Poulton et al., 2012), and an increased incidence of stress fractures in combat military recruits (Schermann et al., 2018). Only a few studies have evaluated the role of this medication in dental health. One retrospective study showed that MP intake did not affect teeth maturation in a group of children who were exposed to the medication for at least two years (Batterson et al., 2005). Regarding oral health (OH), several small observational studies have shown conflicting results (Grooms, Keels, Roberts, & McIver, 2005; Hidas et al., 2011; Rosenberg, Kumar, & Williams, 2014).

The possible association between MP use and OH is confounded by the generally recognized adverse effects of ADHD on OH, outlined by Broadbent *et al*.: 1) lack of motivation to maintain oral hygiene; 2) more frequent reward of children by their parents with cariogenic foods; 3) the medication-related xerostomia and 4) more frequent reporting of caries by guardians and parents of children with ADHD (Broadbent, Ayers, & Thomson, 2004). Significance of the medication-related xerostomia remained unproven. Normal saliva secretion was shown to play a significant role in the prevention of dental caries (Dawes, 2008; Stookey, 2008). However, studies that measured saliva secretion and reported MP use inconclusively argue whether saliva flow is reduced in the MP-treated patients and whether it has a clinically significant effect on caries burden (Friedlander & Friedlander, 1992; Grooms et al., 2005; Hidas et al., 2011).

We suggest that a complex relationship between MP use and oral health exists, including the confounding behavioral effect of ADHD, which may be either intensified or controlled by the medication. This enhances the difficulty to elucidate the effect of the medication alone on oral health and dental treatment needs. Previous small observational studies may have suffered from insufficient power and selection bias, and were inadequate for investigation of the overall effect of MP on oral health.

The goal of the following study was to present a dose-response effect of MP use on restorative treatment needs, which served as an indicator of caries. Data from a large population cohort of young adults were analyzed, while adjusting for multiple factors that are known to affect the number of restorative treatments. We hypothesized that there are higher treatment needs among young adults with ADHD as compared to healthy controls, but no independent association between MP and treatment needs.

## Materials and Methods

After receiving ethical approval and waiver of an informed consent by the IRB, an automated query of the military medical records was performed. The retrospective cohort included all subjects aged 18-25 who served in the Israel Defense Forces (IDF) between 2005 and 2017 for at least 12 months and for a maximum period of service of 4 years, to exclude officers and contract servicemen. Subjects with severe chronic medical conditions or history of recent malignancy who volunteered to service were excluded. Likewise, subjects with missing baseline data (age, weight and height) were regarded as false or error entries in the database, and were therefore excluded.

Based on the diagnosis of ADHD and MP prescription, two study exposure groups were defined: subjects with treated ADHD (at least one MP prescription during the study period) and those with untreated ADHD (no medication prescriptions). The third group consisted of 200,000 of matched healthy controls.

Baseline subject information included age at the time of first encounter at the dental clinic, gender, height (cm), weight (kg), BMI (kg/m^2^), socio-economic status (SES, a measure used by the state Central Bureau of Statistics, that is calculated from mean income of subject’s area of residence and includes 1-10 grades), education level (less than 12 years, 12 years of school and higher education), father’s origin (whether immigrated from Europe and North America, Asia and South America, Mediterranean Africa, Sub-Saharan Africa, former USSR or was Israeli-born) and duration of follow-up in months. SES was categorized to three levels (1-4 low, 5-7 middle, 8-10 high).

Exact MP prescription data throughout the study period was retrieved from patient medical records in order to establish the dose-response effect between MP use and dental treatment needs. Dental treatment data was available through dental clinic records, and included the number of planned restorative treatments per subject. Restorative treatment data included not only actual but also recommended treatments, reflecting number of caries lesions and other restorative treatments required at the same encounter. This allowed estimation of the overall dental treatment needs per subject and was considered more optimal than counting only the actually performed treatments, which could lead to higher confounding by SES. Primary outcome was having a recommendation for at least one restorative treatment. The data did not include prosthodontic and endodontic treatments, and dental extractions. Revision treatments were counted along with primary caries treatments, as no unique treatment code was assigned for these procedures.

### Statistical analysis

Statistical analyses included descriptive statistics, the unadjusted dose-dependent risk estimates, and multivariate logistic regression. Categorical data (education, socioeconomic level and origin) was presented as percentages and was analyzed using the chi-square test. Continuous data (age, height, weight, BMI, fitness test grade and duration of follow-up) was presented as mean ± standard deviations and analyzed using Student’s t-test.

For the purpose of estimating the unadjusted dose-dependent risk of dental treatments, total amount of MP used during the service was summed up. The risk was measured in increments of 1000 mg, which correspond to 3 months of treatment or less. Risk was estimated using linear regression, adjusted only for duration of follow-up.

Finally, a multivariate logistic regression was fitted to adjust the prediction of having at least one restorative treatment for possible confounding variables. Those variables that differed significantly between study groups were included in the model. Relationship between predictors was reviewed to exclude autocorrelation above 0.8. Odds ratios were calculated from the regression coefficients, and their respective confidence intervals were estimated using bootstrapping. Study results were reported according to STROBE guidelines (von Elm et al., 2008). STROBE checklist was submitted along with the manuscript.

## Results

The final cohort consisted of 6,875 subjects with ADHD who received treatment with MP (“Treated”), in addition to 6,729 subjects with ADHD who had no prescriptions for MP (“Untreated”) and 200,000 healthy controls. The subjects’ baseline characteristics are presented in Table 1. Subjects with treated ADHD had an average 3 kg more weight than controls (p<0.001), and more of them belonged to a higher socioeconomic class, compared to both Untreated and Controls (33.2% vs 26.2% vs 21%, p<0.001). Other baseline characteristics were significantly but not substantially different between the groups (Table 1).

**Table 1:** Baseline demographic characteristics of the study subject

| Table 1: Baseline demographic characteristics of the study subject |  |  |  |  |
| --- | --- | --- | --- | --- |
|  | Treated ADHD<br>(n=6,875) | Untreated ADHD<br>(n=6,729) | Healthy controls<br>(n=200,000) | p-value |
| Age | 18.35±0.86 | 18.33±0.8 | 18.4±0.97 (***) | <0.0001 |
| Height | 168.6±11.7 | 169.5±11.5 (***) | 167.9±15.7 (***) | <0.0001 |
| Weight | 66.2±15.3 | 65.7±14.7 (***) | 63.3±14.5 (***) | <0.0001 |
| BMI |  |  |  | <0.0001 |
| <20 | 23% | 27% | 31% |  |
| 20-24 | 51% | 50% | 50% |  |
| 25+ | 26% | 23% | 19% |  |
| % female | 47% | 38% (***) | 44% (***) | <0.0001 |
| Socio-economic level |  |  |  | <0.0001 |
| low | 1.6% | 2.2% | 3% |  |
| middle | 65.2% | 71.6% | 76% |  |
| high | 33.2% | 26.2% | 21% |  |
| Education |  |  |  | <0.0001 |
| <12 years | 5% | 8% | 5% |  |
| 12 years | 92% | 90.5% | 93% |  |
| >12 years | 3% | 1.5% | 2% |  |
| Duration of follow-up<br>(months) | 35±17 | 33±14 | 33±14 | 1 |
| ***Significant at level <0.01 compared to treated ADHD group |  |  |  |  |

The treated group exhibits the highest percent of subjects who required at least one restorative treatment (24%), as well as the highest average number of treatments per person (0.88) (Table 2). On multivariate analysis, the dose-response association between MP use and the odds of having at least one restorative treatment persisted after adjustment for the diagnosis of ADHD and other variables (OR=1.006 for each additional 1000 mg of MP, 95% CI [1.004:1.009]) (Table 3). Additional significant determinants of the need for restorative treatment were extremes of BMI, male gender, lower education level, lower socioeconomic class, older age, longer duration of follow-up and ethnic origin from Sub-Saharan and Northern Africa, compared to subjects whose parents were not immigrants. Subjects from former Soviet Union, as well as subjects from North America and Europe had lower risk of restorative treatments.

**Table 2:** Prevalence of the restorative treatments needs among the study groups

|  |  |  |  |  |
| --- | --- | --- | --- | --- |
|  | Treated ADHD<br>(n=6,875) | Untreated ADHD<br>(n=6,729) | Healthy controls<br>(n=200,000) | p-value |
| % of subjects who required at least one caries treatment | 24% | 22% | 17% | <0.0001 |
| Mean number of caries treatments (mean ± SD) | 0.88 ± 1.69 | 0.78 ± 1.54 | 0.61 ± 1.72 | <0.0001 |

**Table 3:** Determinants of the odds of requiring at least one restorative treatment (Multivariate regression analysis)

|  |  |
| --- | --- |
| Parameter | OR (95%CI) |
| Total of 1000 mg of MP<br>(Risk increasing with each additional 1000 mg MP) | 1.006 (1.004:1.009)*** |
| BMI <20 | 0.97 (0.968:0.978)*** |
| BMI >=25 | 0.988 (0.982:0.994)*** |
| Male gender | 1.04 (1.035:1.045)*** |
| Diagnosis of ADHD | 1.12 (1.11:1.13)*** |
| Education < 12 years | 1.035 (1.02:1.05)*** |
| Education > 12 years | 1.008 (0.99:1.02) |
| Middle socio-economic class | 0.98 (0.97:0.99)** |
| High socio-economic class | 0.96 (0.95:0.97)*** |
| Age | 1.035 (1.033:1.038)*** |
| Months of follow-up | 1.0076 (1.0074:1.0078)*** |
| Origin from Sub-Saharan Africa | 1.04 (1.03:1.06)*** |
| Origin from former USSR | 0.98 (0.97:0.99)*** |
| Origin from North Africa | 1.00175 (1.0017: 1.018)*** |
| Origin from Asia and South America | 1.0018 (0.99:1.01) |
| Origin from Europe and North America | 0.97 (0.97:0.98)*** |
| ***significant at 0.001, ** significant at 0.01<br>OR – Odds Ratio<br>95% CI – 95% Confidence Interval |  |

## Discussion

The study has confirmed the existence of a dose-response effect of MP use on the requirement for restorative treatment in a large cohort of young adults. At first glance, the MP-associated restorative treatments OR of 1.006 for each additional gram of MP may seem negligible. However, it is worth keeping in mind that treated young adults with ADHD typically undergo chronic medication from childhood (Katusic et al., 2005; Mannuzza et al., 2008). Therefore, with treatment duration of years and a total cumulative dose of MP over tens of gr (Katusic et al., 2005; Mannuzza et al., 2008), our results show that chronic MP medication among young adults leads to an elevated need for restorative treatments and implies a significant impact on oral health. Other important findings include the association between restorative and the diagnosis of ADHD, male gender, lower socioeconomic status and education level, extremes of BMI and parents’ immigration from developing countries.

The association between MP use and caries has been questioned by previous studies, but was not supported by evidence yet. Two small retrospective studies have specifically investigated the hypothesis that MP-related xerostomia and loss of teeth-protective salivation may be the mediators of higher prevalence of caries and plaque formation in ADHD children (Rosenberg et al., 2014). Hidas *et al*. have compared oral health status, salivary flow rate and salivary quality across three groups of children, adolescents and young adults: those with ADHD using stimulant medication, those having ADHD but using no medications and healthy controls. When grouped together regardless of the treatment status, subjects with ADHD had lower salivary flow rate, higher plaque index but same decay, missing and filled teeth (DMFT) index as healthy controls. MP use itself apparently had no significant effect on oral health and saliva flow and quality (Hidas et al., 2011). However, the study was limited to about 30 subjects in each group, and may have lacked sufficient power. Grooms *et al*. compared saliva flow and visual dental examination of 38 children with medication-treated ADHD and 38 healthy children. The ADHD group exhibited a higher load of caries lesions. At the same time, amount of saliva, fluoride exposure, dental floss use and diet were similar between the groups (Grooms et al., 2005). Following the above research, a recent meta-analysis has concluded that decreased salivary flow does not appear to be responsible for higher prevalence of caries in ADHD children (Rosenberg et al., 2014). This conclusion had left the question open and demanded a re-evaluation of caries prevalence among MP-treated ADHD children.

In the present study, we were able to reliably estimate the overall burden of caries in a large sample of military recruits, represented by restorative treatment needs. Due to the compulsory conscription law, all population groups were adequately presented in the study sample and can be generalized to a respective population of the same age in a developed country. All subjects had equal access to the military dental service, resulting in accurate documentation of the recommended treatments. The study subjects could choose the alternative private dental practices, but we estimate that the study results and conclusions were not affected based on the following reasons. First, the study was based on records of recommended treatments. It is plausible that the subjects who eventually underwent treatment in private clinics, had initially underwent assessment in military clinics and received a treatment plan. Second, this sort of bias would have affected subjects with higher socioeconomic status to a larger extend than it would affect those from less favorable economic background, i.e. would lead to underestimation of caries rates among the former. However, there were more subjects with higher socioeconomic status in the group of MP-treated ADHD subjects, who had highest treatment rates despite the possible underestimation.

One of the strengths of this study is the multivariate adjustment of the association between MP use and caries restorative treatments to several important upstream risk factors for caries. These include lower income (André Kramer, Pivodic, Hakeberg, & Östberg, 2019; Baelum, 2011) and education level (Arrica et al., 2017; Carta et al., 2015). This study confirms previous findings among IDF recruits of an inverse association between the socioeconomic status and the number of restorative treatments (Levy, Livny, Sgan-Cohen, & Yavnai, 2018). Interestingly, the subjects immigration status could have either protective or adverse effect on restorative treatment needs. This may be partially explained by the fact that immigration to Israel originates from various countries; immigrants from developed and developing countries accordingly incorporate in higher and lower socioeconomic communities in Israel. Immigrants originating from Ethiopia had higher restorative treatment needs compared to non-immigrants, in agreement with previous publications (Levy et al., 2018; Sgan-Cohen, Katz, Horev, Dinte, & Eldad, 2000). Origin from former USSR was associated with lower risk of restorative treatment need in this study, contrary to previous findings of research among young Israeli adults (Birnboim-Blau, Levin, & Sgan-Cohen, 2006; Sgan-Cohen et al., 2000). However, the disparity is non-significant: one of these studies focused on combat soldiers only and showed a negligible increase in restorative treatment needs (RR = 1.074, p<0.001) (Levy et al., 2018), whereas other two were conducted in a different time period (Birnboim-Blau et al., 2006; Sgan-Cohen et al., 2000).

The limitations of this study are retrospective design and indirect assessment of the caries burden (number of recommended treatments was used instead of standard measures used in prospective studies). In addition, saliva secretion could not be assessed in this study, and its role in the pathophysiology of the effect of MP on caries incidence remains obscure. Outcome data had probably overestimated caries incidence, because the code for “preservation treatments” had also included restoration of broken cements and teeth, and not only active caries lesions. On the other hand, it did not account for severe lesions that required root treatment or crown placement. Higher amount of treatments in MP has probably been confounded by socioeconomic status. There were significantly more subjects from higher socioeconomic status among MP users, who were more likely to schedule encounter at a dental clinic. Moreover, higher utilization of services could lead to higher number of treatment recommendations, creating a self-sustaining cycle. On the other hand, this effect may be mitigated given that these subjects were also more likely to turn to private dental services outside the military dental service.

This study represents the first ecological evidence of a dose-response effect of MP use and incidence of restorative treatment needs. Subjects with ADHD who receive chronic treatment with MP have higher restorative treatment needs than subjects with untreated ADHD and healthy controls. Our results show that chronic MP medication among young adults leads to an elevated need for restorative treatments and implies a significant impact on oral health. The pathophysiology behind this association remains uncharacterized and prompts for further investigation.

## Data Availability

Not available

## Acknowledgments

None

## Statement of Ethics

The study protocol has been approved by the research institute’s institutional review board for human research. The requirement for informed consent was waived by the committee.

---

**Figure 1:**
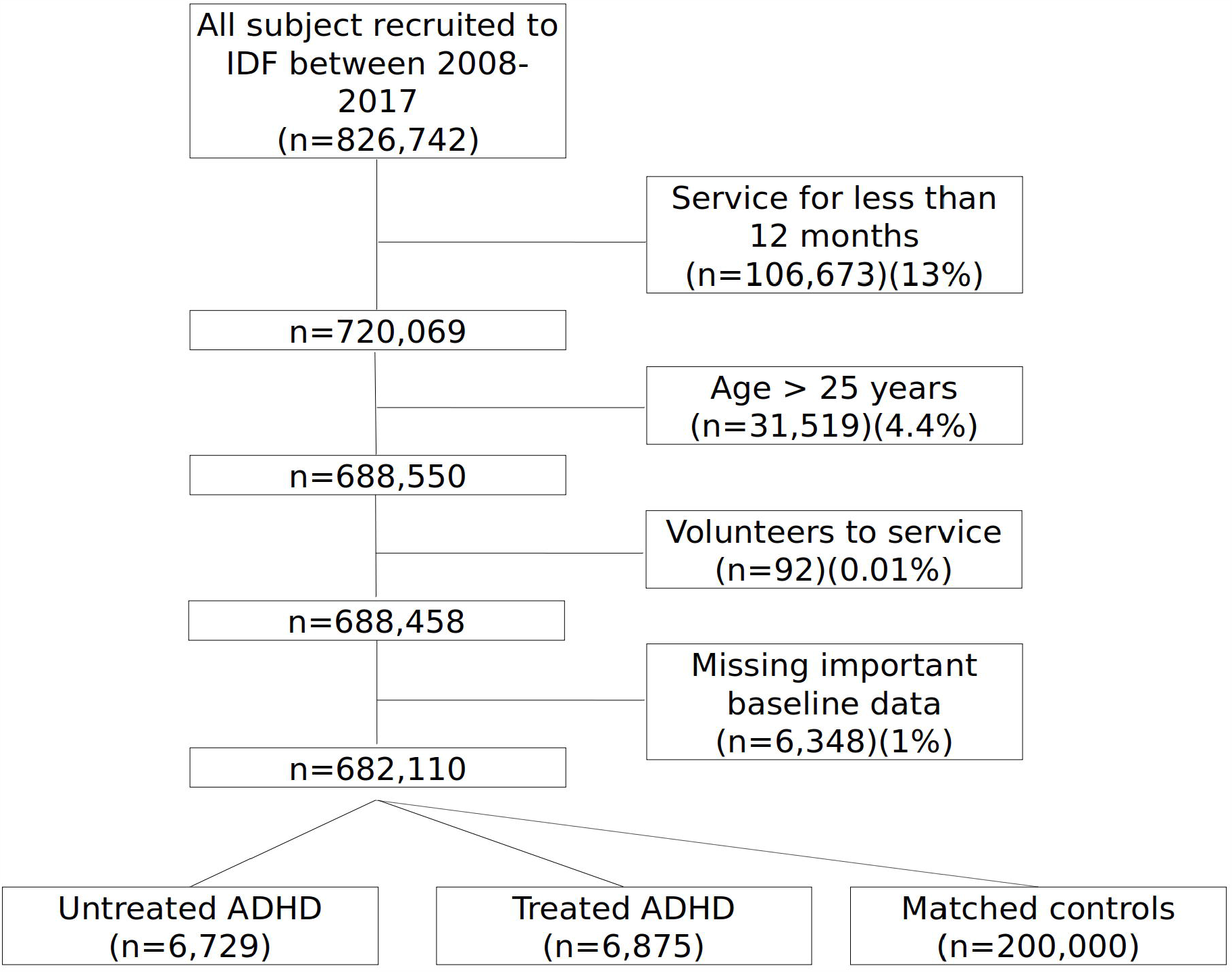
Study flowchart

